# COVerAGE-DB: A database of age-structured COVID-19 cases and deaths

**DOI:** 10.1101/2020.09.18.20197228

**Authors:** Tim Riffe, Enrique Acosta, COVerAGE-DB project team, Jose Manuel Aburto, Diego Alburez-Gutierrez, Anna Altová, Ugofilippo Basellini, Simona Bignami, Didier Breton, Eungang Choi, Jorge Cimentada, Gonzalo De Armas, Emanuele Del Fava, Alicia Delgado, Viorela Diaconu, Jessica Donzowa, Christian Dudel, Antonia Fröhlich, Alain Gagnon, Mariana Garcia Cristómo, Victor M. Garcia-Guerrero, Armando González-Díaz, Irwin Hecker, Dagnon Eric Koba, Marina Kolobova, Mine Kühn, Mélanie Lépori, Chia Liu, Andrea Lozer, Madalina Manea, Muntasir Masum, Ryohei Mogi, Celine Monicolle, Saskia Morwinsky, Ronald Musizvingoza, Mikko Myrskylä, Marilia R. Nepomuceno, Michelle Nickel, Natalie Nitsche, Anna Oksuzyan, Samuel Oladele, Emmanuel Olamijuwon, Oluwafunke Omodara, Soumaila Ouedraogo, Mariana Paredes, Marius Pascariu, Manuel Piriz, Raquel Pollero, Larbi Qanni, Federico Rehermann, Filipe Ribeiro, Silvia Rizzi, Francisco Rowe, Isaac Sasson, Jiaxin Shi, Rafael Silva-Ramirez, Cosmo Strozza, Catalina Torres, Sergi Trias-Llimos, Fumiya Uchikoshi, Alyson van Raalte, Paola Vazquez-Castillo, Estevão Vilela, Iván Williams, Virginia Zarulli

**Author notes:** co-first author (Tim Riffe, Enrique Acosta). The full list of authors and contributions within the COVerAGE-DB project are presented at the end of the manuscript. **Suggested display authors:** Tim Riffe, Enrique Acosta, and the COVerAGE-DB team.

## Abstract

COVerAGE-DB is an open-access database including cumulative counts of confirmed COVID-19 cases, deaths, and tests by age and sex. The main goal of COVerAGE-DB is to provide a centralized, standardized, age-harmonized, and fully reproducible database of COVID-19 data. Original data and sources are provided alongside data and measures in age-harmonized formats. An international team, composed of more than 60 researchers, contributed to the collection of data and metadata in COVerAGE-DB from governmental institutions, as well as to the design and implementation of the data processing and validation pipeline. The database is still in development, and at this writing, it includes 89 countries, and 237 subnational areas. Cumulative counts of COVID-19 cases, deaths, and tests are recorded daily (when possible) since January 2020. Many time series thus fully capture the first pandemic wave and the beginning of later waves. Since collection efforts began for COVerAGE-DB several studies have used the data.

## Data Resource Basics

Information about pandemic dynamics is critical to understand the potential impacts on populations, design mitigation strategies, and evaluate the efficacy of their implementation. Centralization, standardization, and harmonization of data is critical to enable comparisons of the demographic impact of COVID-19 vis-à-vis differences in the age-compositions of confirmed infections and deaths. The international data landscape must keep pace with the global march of the pandemic, and researchers must work to triangulate the available data to create comparable measures to monitor and predict its demographic impacts.

The COVID Age Database (COVerAGE-DB) aims to provide global coverage of key demographic aspects of the COVID-19 pandemic as it unfolds in an up-to-date, transparent, and open-access format. COVerAGE-DB offers data with standardized count measures and harmonized age groups to allow comparisons between populations at national and subnational scales.

The database is currently under expansion through both the increase in coverage of national and subnational populations and the inclusion of more recent periods as the pandemic continues to unfold. At this writing, the database contains daily counts of COVID-19 cases, deaths, and tests performed by age and sex for **89** national and **237** subnational populations around the world, depending on the available data for each source. The date range available for each country or subpopulation varies. In several country series, the database includes the earliest confirmed cases in January, 2020. For most populations the database includes daily time series, beginning from an initial starting date when the data were first released or collected by our team. Fig. 1 displays a map of countries included in the database, indicating at least one subnational population from 12 countries. A detailed overview of data availability is given in a searchable table [external link](https://bit.ly/3kVDrLD)

**Figure 1.**
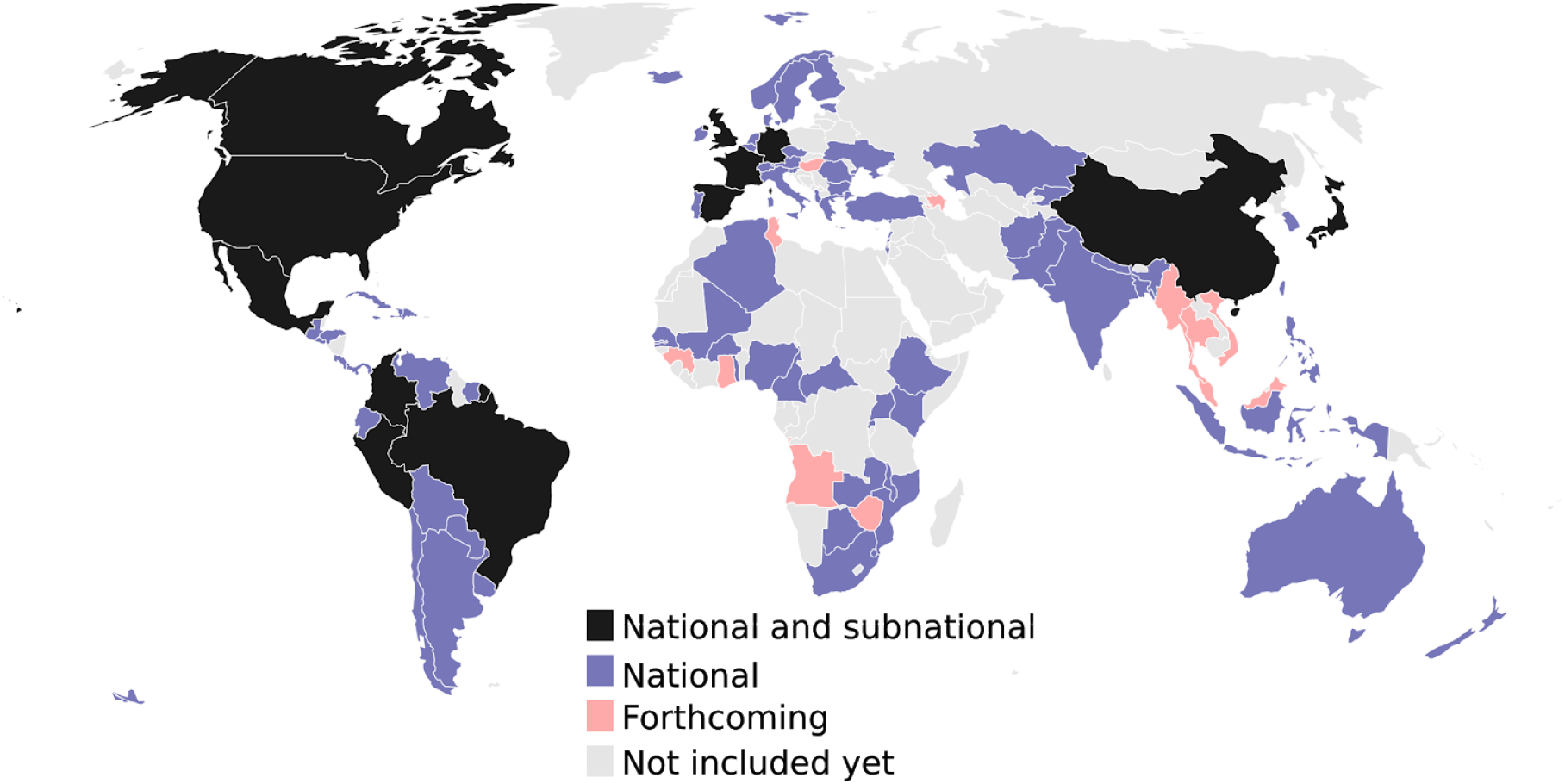
Availability of national- and subnational information on COVID-19 cases, deaths, and tests in the countries included in the database as of 24 August 2020.

### Data Collected

#### Collection

Official counts of COVID-19 cases, deaths, and tests are extracted from reports published by official governmental institutions, such as health ministries and statistical offices. Depending on the source, data are collected in a variety of formats, including machine-readable files, pdf tables, html tables, interactive dashboards, press releases, official announcements via Twitter, and in a few instances, from digitized graphics. A full list of data sources is available in a dashboard view (https://bit.ly/2Qg1MxL).

Generally, COVID-19 cases, deaths and tests are reported as counts in 10-year age groups, but some sources report data in other metrics (fractions, percents, ratios) or as summary indicators such as case fatality ratios by age. Reported age intervals vary by source, ranging from single ages to 30-year or greater age bands, and sometimes reported age intervals change over time within sources. Usually data are reported as cross-sectional snapshots of cumulative counts, but some sources give full time series of new cases or deaths, in which case we cumulate counts over time. We also collect standard metadata on each of the sources to capture various characteristics of the collected data, such as the primary collection channels, definitions used, and notes on major disruptions or events. An overview of key fields from this metadata is shared as a spreadsheet (https://bit.ly/2FAmKFn).

#### Production

All source data is entered into standard spreadsheet templates hosted in a central folder on Google Drive. Data entry into the templates is either manual or automatic depending on the source.

R programs collect data from the source templates and compile the merged input database. The merged input file is then subject to a series of automatic validity checks. Initial checks are carried out by the individual responsible for data collection and entry using an interactive application (https://mpidr.shinyapps.io/cleaning_tracker/). Data is then harmonized to standard metrics (counts), measures (cases, deaths, tests), and age bands (5- and 10-year age intervals). Harmonization procedures include various kinds of rescaling to ensure coherence in marginal sums. Age group harmonization is done using the penalized composite link model,^1^ which was designed for splitting histograms of count data.

The complete details on all steps of production are available in the COVerAGE-DB Method Protocol, which is publicly available on the web (https://osf.io/jcnw3/). A table listing which adjustments are applied to each population is available on the project website (https://bit.ly/2E61BSV). Both the merged input database and the harmonized output files are uploaded daily as zipped csv files to an Open Science Framework repository (OSF) (https://osf.io/mpwjq/). A GitHub repository (https://bit.ly/2YbtPCJ), which is linked to OSF, contains all R scripts used in the complete production pipeline, including compilation, diagnostics, and harmonization.

### Data Resource Use

Since collection efforts began for COVerAGE-DB in late March 2020, we are aware of 6 studies using the data, many of which provide R code online and are fully reproducible. These studies aim to: (i) explore country differences in the age distribution of COVID-19 deaths,^2^ (ii) assess the contribution of infection case age-structure and age-specific fatality to between- and within-country differences in case fatality rates (CFR) associated with the COVID-19,^3^ (iii) produce a standard age-specific case fatality rate pattern for an indirect demographic method to estimate COVID-19 total infections,^4^ (iv) analyze the association between intergenerational relationships and COVID-19 fatality rates,^5^ (v) estimate years of life lost due to Covid-19,^6^ and (vi) calculate pooled sex ratios of age-specific CFRs of COVID-19 in Europe.^7^ The database is also used to monitor COVID-19 impacts in particular age ranges, for instance, UNICEF uses the database for monitoring the burden of the pandemic on the infant, child, and juvenile mortality around the world.

As an example of the analyses that COVerAGE-DB enables, Fig. 2 displays changes in the relation between age-specific deaths and cases in Colombia, inspired by Fig. 1 of Dudel et al.^3^ We divide both cases and deaths in each age band by the respective population sizes. Diagonal lines indicate implied age-specific case fatality rates. The graphic illustrates a sharp increase in CFR over age for each sex, and it also displays considerable sex differences. For instance, men aged 60-69 in Colombia have almost the same case fatality rates (approximately 12% risk of death after COVID-19 infection is diagnosed) as women aged 70-79.

**Figure 2.**
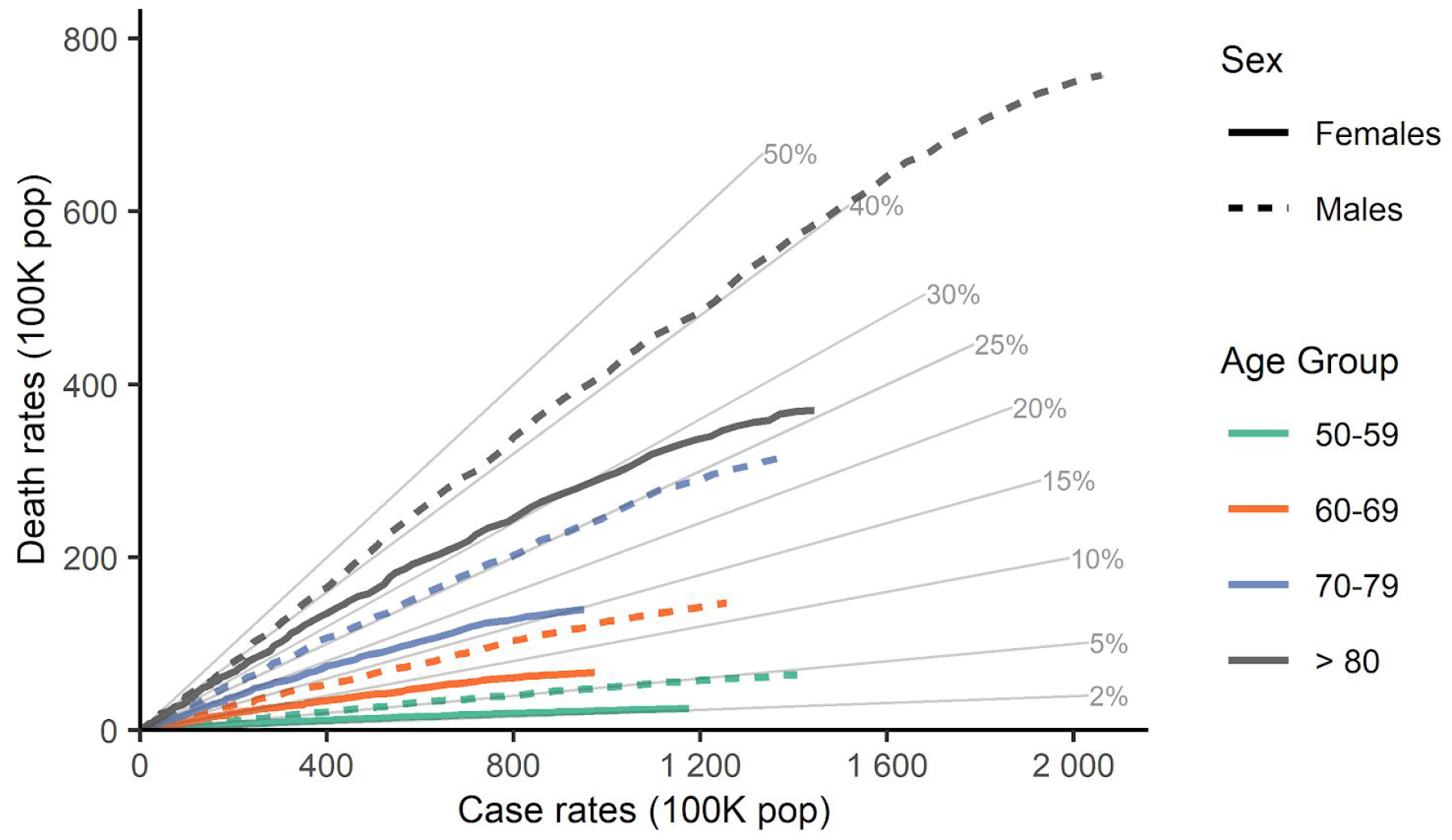
Relation between deaths and cases per 100,000 population by age group and sex in Colombia, until 22 August 2020. Diagonal lines indicate the case fatality rate.

We repeat this exercise to compare Colombia with Mexico (see Fig. 3), where standardizing by population size is more justified. Case fatality rates and death rates are much higher in Mexico than in Colombia in each age band – around 2-fold -, except for ages 80+, which show a substantial reduction in the case fatality rate difference, and much higher death rates for Colombia. While we cannot separate the reasons for this with precision, it is clear that Colombia has conducted about twice as many tests per positive case than has Mexico, and positivity trends have been on the rise in both countries.^8^ However, we do not know the age pattern of positivity in these two countries. On the other hand, both countries have excellent open data practices, allowing for construction of detailed series taking into account retrospective corrections. This highlights challenges in making such comparisons, but also the need to produce data with sufficient detail to adjust for biases. It is our view that researchers should triangulate creatively from all available data rather than avoiding difficult comparisons.

**Figure 3.**
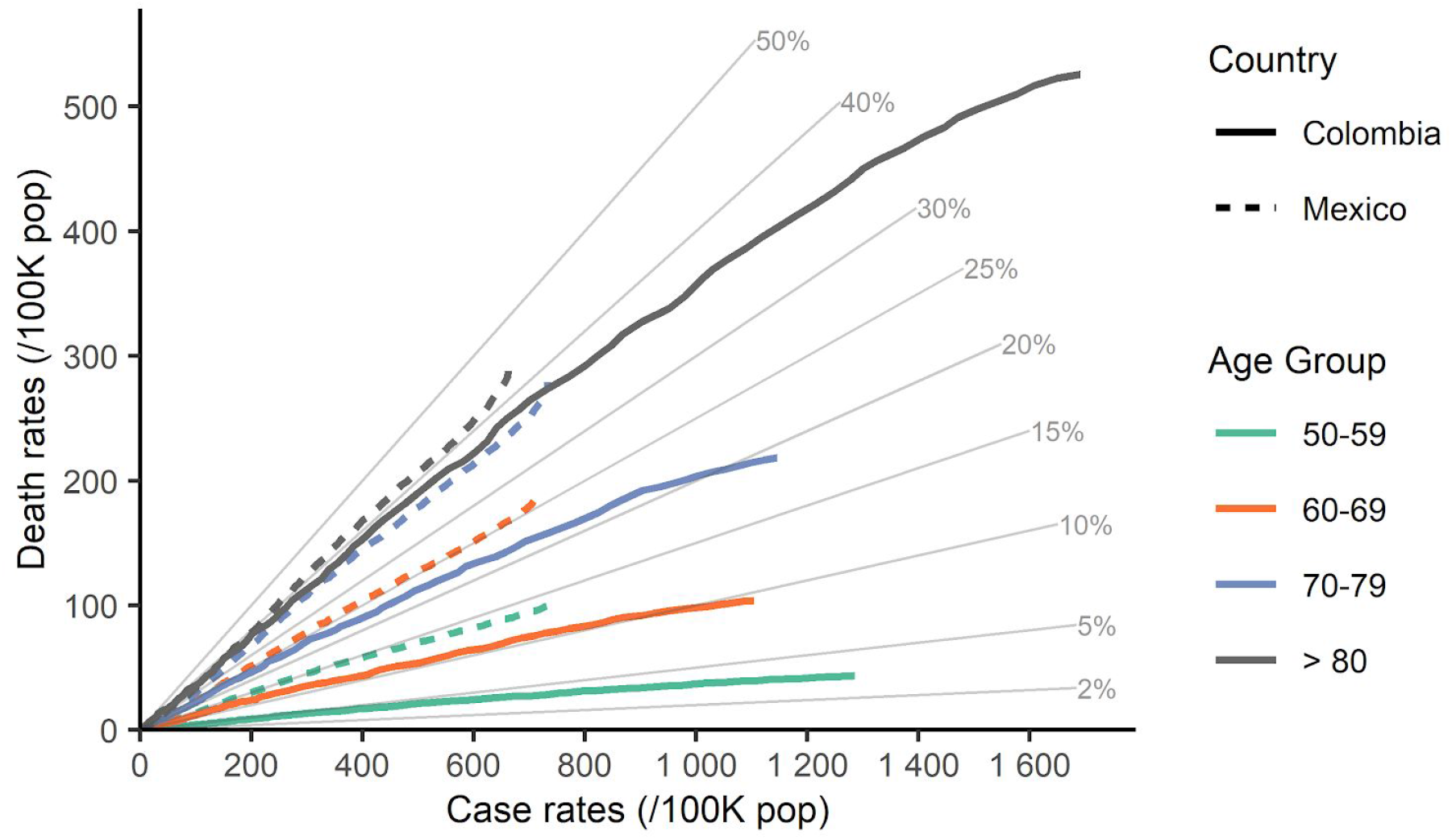
Relation between deaths and cases per 100,000 population by age group in Mexico and Colombia, until 22 August 2020. Diagonal lines indicate the case fatality rate.

## Strengths and Weaknesses

Since the beginning of the pandemic, it has been evident that population characteristics are key to understanding the prevalence, spread and fatality of COVID-19 across countries. However, data on cases, deaths, and tests disaggregated by age and sex are not easily comparable across countries, and sometimes not even accessible. The main strength of COVerAGE-DB is to provide a centralized, open-access, and fully reproducible repository of age-and sex-specific case, death, and test counts from COVID-19, collected from official sources, and harmonized to standard output formats. The data harmonization process is transparent, following a strict protocol.^9^ The initial input data is provided alongside the harmonized counts, as well as the code used to harmonize the different input measures, metrics, and age groups into comparable granular output metrics. All scripts are written in the open-source R programming language.^10^ The data sources and limitations are documented for each country in a standard metadata framework.

A limitation of the COVerAGE-DB is the heterogeneous and difficult-to-evaluate quality of the underlying data. No single data source can currently claim accurate estimates of COVID-19 incidence or fatalities. Age-specific case counts are highly dependent upon the testing capacity,^11^ testing strategy,^12^ and differences in the definition of cases across sources and over time. Cases are underestimated everywhere, with underestimation expected to vary by age, given the relationship between age and case severity.^13^ The accuracy of diagnostic RT-PCR tests used to confirm infections is also known to vary.^14^ Furthermore, at any given date, cumulative counts are underestimated because of the lag between infection and a positive test result.^15^

Death counts from COVID-19 are also likely underestimated for similar reasons, but also due to various kinds of delays in death registration. Media reports have circulated about intentional data manipulation in some of the official data covered in the database.^16^ Excess all-cause mortality has been observed across many regions.^17–20^ Although some of these deaths are likely from postponing or foregoing treatment from non-COVID-19 related causes, the magnitude of this excess is suggestive that numerous COVID-19 related deaths are classified under different causes. Populations also differ in whether deaths to suspected COVID-19 cases are included in official statistics and in post-mortem practices when an infection is suspected.^21^ Some populations only report deaths occurring in hospitals, neglecting a potentially sizable proportion of deaths occurring in institutional settings and at home.^22^ While most populations currently report all deaths to confirmed COVID-19 infections as COVID-19 deaths for this database, the underlying cause of death eventually reported on death certificates may differ in patients with severe comorbidities. To mitigate biases and misinterpretations due to different practices and definitions, such information is constantly updated and documented in the metadata of the database, which is freely accessible to users.

All of these issues compromise the comparability of the data contained within the COVerAGE-DB, both across populations at any given time and within populations over time. For these reasons, infection fatality ratios, which include in the numerator detected and undetected COVID-19 infections, should not be estimated from COVerAGE-DB data alone. Proper estimation of incidence and fatality will likely require triangulating data across numerous sources as these become available. To this end, the COVerAGE-DB was designed to be easily merged with other databases such as the Our World In Data database on COVID-19 testing,^8^ the COVID-19 dashboard of Johns Hopkins,^23^ the World Population Prospects database,^24^ and the Short Term Mortality Fluctuations database.^20^ Moreover, given that we have near-complete time series capturing the whole pandemic curve in some places, careful modeling of the lag structures might allow some of these data-driven biases to be estimated.

## Data Resource Access

Both merged input and harmonized output files can be downloaded directly from the OSF site (https://osf.io/mpwjq, DOI: 10.17605/OSF.IO/MPWJQ), which contains a folder called *Data* with three files of primary data. Fig 4 shows where to find the files in the OSF repository.

**Figure 4.**
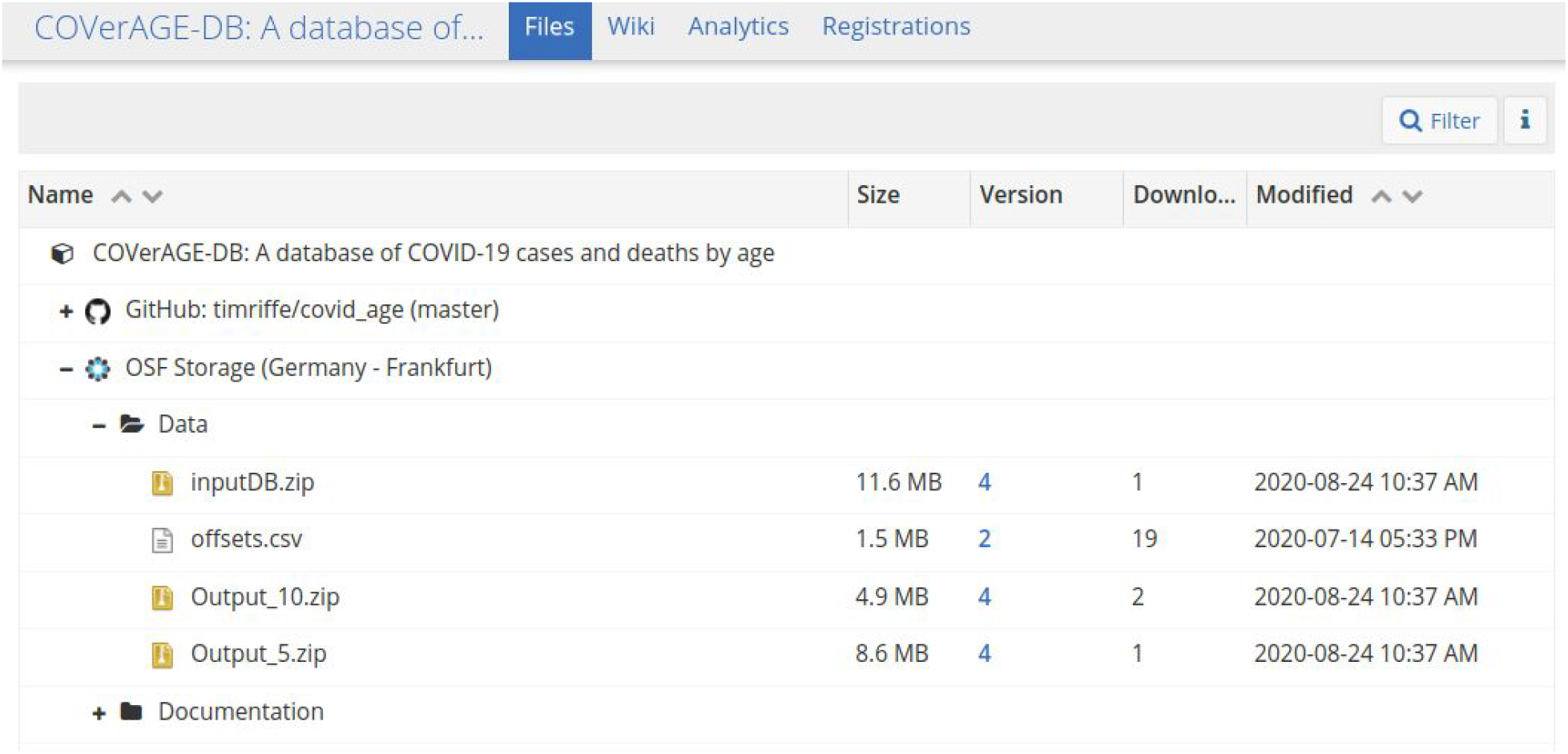
View of the Open Science Framework (OSF) repository, File section (https://osf.io/mpwjq/files/). To download data files, click on `Data`, and select one of the files.

Each of the main data files has a stable link (see Tab. 1), which always points to the most recent version. Each file is a zipped csv file by the same name.

**Table 1.**
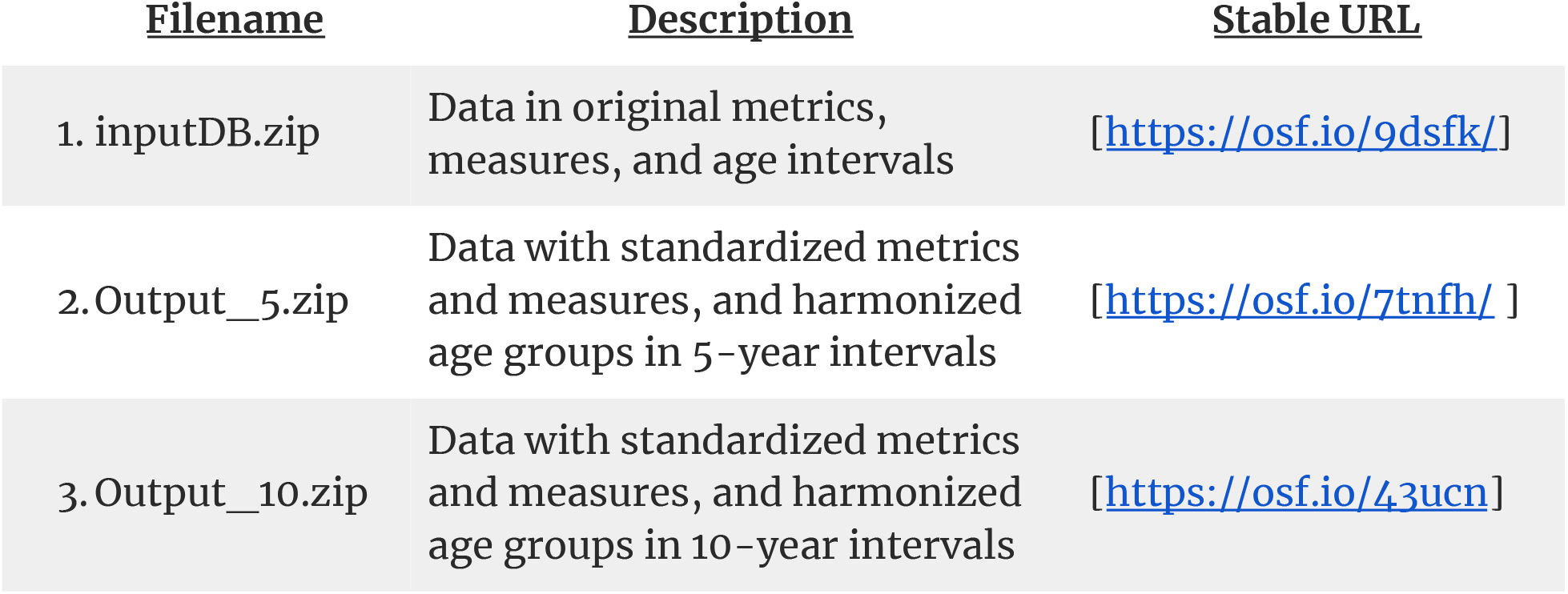
The main data files, a description of their content, and their stable URLs.

| <u>Filename</u> | <u>Description</u> | <u>Stable URL</u> |
| --- | --- | --- |
| 1. inputDB.zip | Data in original metrics, measures, and age intervals | [https://osf.io/9dsfk/] |
| 2. Output_5.zip | Data with standardized metrics and measures, and harmonized age groups in 5-year intervals | [https://osf.io/7tnfh/] |
| 3. Output_10.zip | Data with standardized metrics and measures, and harmonized age groups in 10-year intervals | [https://osf.io/43ucn/] |

For stable links to download particular versions, click on the version number in the *Version* column seen in Fig. 4. Users can note versions either by referring to timestamps provided in the headers of data files or by referring to OSF file version numbers, which increment with each daily update.

A data dictionary is given in both the OSF wiki (https://osf.io/mpwjq/wiki/home/) and the Method Protocol. Files are shared in csv format to be as universally accessible as possible. A guide to getting started using the data in R is also provided (https://bit.ly/3g8nIVU), and tips for other statistical packages may also be added. Users are encouraged to reach out for further information or advice on using the database, or to express interest in the project.

## Data Availability

All data are openly available and free in an OSF repository. All processing code is openly available and free in a github repository.

https://osf.io/mpwjq/

https://github.com/timriffe/covid_age

## Acknowledgments

We gratefully acknowledge the hard work of health ministries and statistical offices around the world in preparing and disseminating the data included in COVerAGE-DB.

## Notes

The COVerAGE-DB project team is composed of more than 60 researchers that share in the authorship of this manuscript according to the CRediT authorship system (see contributions by author under the following link: https://docs.google.com/spreadsheets/d/1SHygPJArbfInXlmFku6vR7LzLyGz00p1ZkBaPdanAIw/edit?usp=sharing).

**Last, First Affiliation**

Riffe, Tim Max Planck Institute for Demographic Research, Germany

Acosta, Enrique Max Planck Institute for Demographic Research, Germany

Aburto, José Manuel University of Oxford, UK

Alburez-Gutierrez, Diego Max Planck Institute for Demographic Research, Germany

Altová, Anna Faculty of Science, Charles University, Czechia

Basellini, Ugofilippo Max Planck Institute for Demographic Research, Germany

Bignami, Simona University of Montreal, Canada

Breton, Didier Université de Strasbourg, France

Choi, Eungang Ohio State University, USA

Cimentada, Jorge Max Planck Institute for Demographic Research, Germany

De Armas, Gonzalo Universidad de la República, Uruguay

Del Fava, Emanuele Max Planck Institute for Demographic Research, Germany

Delgado, Alicia Pontificia Universidad Católica del Ecuador, Ecuador

Diaconu, Viorela Max Planck Institute for Demographic Research, Germany

Donzowa, Jessica Max Planck Institute for Demographic Research, Germany

Dudel, Christian Max Planck Institute for Demographic Research, Germany

Fröhlich, Antonia Max Planck Institute for Demographic Research, Germany

Gagnon, Alain University of Montreal, Canada

Garcia Cristómo, Mariana El Colegio de México, México

Garcia-Guerrero, Victor M. El Colegio de México, México

González-Díaz, Armando El Colegio de México, México

Hecker, Irwin Université de Strasbourg, France

Koba, Dagnon Eric L’Agence Française de Développement, France

Kolobova, Marina Max Planck Institute for Demographic Research, Germany

Kühn, Mine Max Planck Institute for Demographic Research, Germany

Lépori, Mélanie Université de Strasbourg, France

Liu, Chia St. Andrews University, UK

Lozer, Andrea Max Planck Institute for Demographic Research, Germany

Manea, Mădălina Research Institute for the Quality of Life, Romania

Masum, Muntasir Univ. Texas San Antonio, USA

Mogi, Ryohei Centre for Demographic Studies, Spain

Monicolle, Celine Université de Strasbourg, France

Morwinsky, Saskia Max Planck Institute for Demographic Research, Germany

Musizvingoza, Ronald Bursa Uludag University, Turkey

Myrskylä,Mikko Max Planck Institute for Demographic Research, Germany

Nepomuceno, Marília R. Max Planck Institute for Demographic Research, Germany

Nickel, Michelle Max Planck Institute for Demographic Research, Germany

Nitsche, Natalie Max Planck Institute for Demographic Research,

Germany

Oksuzyan, Anna Max Planck Institute for Demographic Research, Germany

Oladele, Samuel Federal University Oye Ekiti, Nigeria

Olamijuwon, Emmanuel University of the Witwatersrand, South Africa

Omodara, Oluwafunke Federal University Oye Ekiti, Nigeria

Ouedraogo, Soumaila French Institute for Demographic Studies, France

Paredes, Mariana Universidad de la República, Uruguay

Pascariu, Marius SCOR, France

Piriz, Manuel Universidad de la República, Uruguay

Pollero, Raquel Universidad de la República, Uruguay

Qanni, Larbi Max Planck Institute for Demographic Research, Germany

Rehermann, Federico Universidad de la República, Uruguay

Ribeiro, Filipe CIDEHUS, Universidade de Évora, Portugal

Rizzi, Silvia University of Southern Denmark, Denmark

Rowe, Francisco University of Liverpool, UK

Sasson, Isaac Tel Aviv University, Israel

Shi, Jiaxin Max Planck Institute for Demographic Research, Germany

Silva-Ramirez, Rafael University of Montreal, Canada

Strozza, Cosmo University of Southern Denmark, Denmark

Torres, Catalina Muséum national d’histoire naturelle, France

Trias-Llimos, Sergi Centre for Demographic Studies, Spain

Uchikoshi, Fumiya Princeton University, USA

van Raalte, Alyson Max Planck Institute for Demographic Research, Germany

Vazquez-Castillo, Paola El Colegio de México, México

Vilela, Estevão Universidade Federal de Minas Gerais, Brazil

Williams, Iván Universidad de Buenos Aires / INDEC, Argentina

Zarulli, Virginia University of Southern Denmark, Denmark

